# High-Dimensional Single-Cell Multimodal Landscape of Human Carotid Atherosclerosis

**DOI:** 10.1101/2023.07.13.23292633

**Authors:** Alexander C. Bashore, Hanying Yan, Chenyi Xue, Lucie Y. Zhu, Eunyoung Kim, Thomas Mawson, Johana Coronel, Allen Chung, Sebastian Ho, Leila S. Ross, Michael Kissner, Emmanuelle Passegué, Robert C. Bauer, Lars Maegdefessel, Mingyao Li, Muredach P. Reilly

## Abstract

**Background:** Atherosclerotic plaques are complex tissues composed of a heterogeneous mixture of cells. However, we have limited understanding of the comprehensive transcriptional and phenotypical landscape of the cells within these lesions.

**Methods:** To characterize the landscape of human carotid atherosclerosis in greater detail, we combined cellular indexing of transcriptomes and epitopes by sequencing (CITE-seq) and single-cell RNA sequencing (scRNA-seq) to classify all cell types within lesions (n=21; 13 symptomatic) to achieve a comprehensive multimodal understanding of the cellular identities of atherosclerosis and their association with clinical pathophysiology.

**Results:** We identified 25 distinct cell populations each having a unique multi-omic signature, including macrophages, T cells, NK cells, mast cells, B cells, plasma cells, neutrophils, dendritic cells, endothelial cells, fibroblasts, and smooth muscle cells (SMCs). Within the macrophage populations, we identified 2 proinflammatory subsets that were enriched in IL1B or C1Q expression, 2 distinct TREM2 positive foam cell subsets, one of which also expressed inflammatory genes, as well as subpopulations displaying a proliferative gene expression signature and one expressing SMC-specific genes and upregulation of fibrotic pathways. An in-depth characterization uncovered several subsets of SMCs and fibroblasts, including a SMC-derived foam cell. We localized this foamy SMC to the deep intima of coronary atherosclerotic lesions. Using CITE-seq data, we also developed the first flow cytometry panel, using cell surface proteins CD29, CD142, and CD90, to isolate SMC-derived cells from lesions. Last, we found that the proportion of efferocytotic macrophages, classically activated endothelial cells, contractile and modulated SMC-derived cell types were reduced, and inflammatory SMCs were enriched in plaques of clinically symptomatic vs. asymptomatic patients.

**Conclusions:** Our multimodal atlas of cell populations within atherosclerosis provides novel insights into the diversity, phenotype, location, isolation, and clinical relevance of the unique cellular composition of human carotid atherosclerosis. This facilitates both the mapping of cardiovascular disease susceptibility loci to specific cell types as well as the identification of novel molecular and cellular therapeutic targets for treatment of the disease.

## Introduction

Atherosclerosis, the major underlying cause of cardiovascular diseases (CVD), is a chronic inflammatory disease that is initiated by the infiltration and modification of circulating low-density lipoproteins into the subendothelial space. As the disease initiates and progresses, continuous recruitment of circulating immune cells contributes to a complex inflammatory microenvironment. Additionally, smooth muscle cells (SMCs) migrate from the arterial media into the developing lesion, where they undergo phenotypic modulation and transition to a variety of cell fates including foam cells, macrophage-like cells, synthetic extracellular matrix-producing fibrotic cells, and osteogenic-like cells. Emerging genetic and experimental data suggest that specific SMC-derived cell types are detrimental and contribute to plaque instability (e.g., macrophage and osteogenic types), whereas others are protective and contribute to plaque stability (e.g., fibrotic cell types)^1^. Increased proportions of pro-inflammatory cells relative to matrix-producing cells within the lesion appear to increase plaque vulnerability and the likelihood of rupture, leading to clinical manifestations such as myocardial infarction and ischemic stroke^2^.

Histological studies have identified culprit lesions as having a thin fibrous cap, large necrotic core, and an overabundance of immune cells like macrophages and T cells^3, 4^. Moreover, recent genome-wide association studies (GWAS) have identified many genetic loci associated with CVD^5^, and many of these have been mapped to genes that modulate disease specifically through SMC-derived cell, endothelial cell (EC), or immune cell functions. However, these discoveries have translated into new therapies since the cellular mechanisms driving this complex process have yet to be elucidated fully, cell and molecular mechanistic and causal studies are lacking for most loci, and because mechanistic mouse models may not fully recapitulate the complexity of human atherosclerosis^6^.

Recently, there has been an effort to characterize all cell types in human atherosclerosis by utilizing rapidly evolving single-cell technologies to identify cell type-specific candidate genes and molecular mechanisms and to drive novel therapies for atherosclerosis. These studies have relied primarily on single-cell RNA sequencing (scRNA-seq), which uncovers cellular heterogeneity by identifying subpopulations of cells with distinct transcriptional profiles in atherosclerotic plaques. In one study, a multi-omic approach identified innate and adaptive immune cell alterations in carotid plaques associated with clinical events^8^. In another important study, a vast heterogeneity of all cell types was revealed using combined scRNA-seq and single-cell ATAC sequencing to profile immune and nonimmune cells in human carotid atherosclerosis^9^. To date, these studies have been relatively small in scale and have not provided a precise reference of cell types and canonical protein markers to facilitate clinical and therapeutic translation. To achieve greater precision, we performed CITE-seq utilizing a large panel of antibodies on carotid atherosclerotic plaques in combination with scRNA-seq. Here we report the most comprehensive phenotypic and transcriptional landscape of all cell types within advanced carotid atherosclerosis and identify cell-type specific protein markers as well as perturbed activation states associated with cardiovascular events in the largest set of human samples to date.

## Methods

### Human studies

All human subjects research in this study, including the use of human tissues, conformed to the principles outlined in the Declaration of Helsinki. All patient information was de-identified. For single-cell studies, tissues from human carotid atherosclerotic plaques were collected from twenty-one patients undergoing carotid endarterectomy surgery. These human subject studies were performed with approval (protocol number AAAJ2765) of the local Institutional Review Board (IRB) of Columbia University Irving Medical Center, and written informed consent was obtained from all participants. Exclusion criteria include current infection, known immune system disorder, and active or recent (within last three months) radiation, chemotherapy, hormone-based, and/or immunotherapy treatment for cancer. **Table S1** summarizes the demographic and clinical characteristics of the entire cohort, and subgroups to perform comparative analysis. Symptomatic patients were defined as having a stroke or transient ischemic attack (TIA) within 6 months of surgery. Asymptomatic patients had no history of ischemic events. For arterial immunohistochemistry, human coronary artery tissues were obtained from either donor hearts rejected for transplantation or explanted hearts during cardiac transplant surgery. These human subject studies were performed with approval (protocol number AAAR6796) of the local Institutional Review Board (IRB) of Columbia University. Written informed consent was obtained upon admission before surgery.

### Atherosclerotic plaque cell dissociation for single-cell RNA-seq and CITE-seq preparation

Plaque specimens at endarterectomy were placed in phosphate buffered saline (PBS), immediately transported to the laboratory, washed three times in PBS and minced into small pieces. The tissue was then digested in RPMI 1640 containing Liberase (10 U/mL, Sigma cat no 5401127001), Elastase (8 U/mL, Sigma cat no E7885-20MG), and DNAse (120 U/mL, Worthington cat no LS006331) at 37°C for 1 hour. The cell suspension was filtered through a 70-μm cell strainer, washed with double the volume of RPMI 1640 with 10% FBS, centrifuged at 400g for 5 min at 4°C, washed once with sterile-filtered FACS buffer (PBS + 2% FBS + 5mM EDTA pH 8.0 + 20mM HEPES + 1mM sodium pyruvate), and spun down at 400g for 5 min at 4°C. For CITE-seq, plaque specimens were dissociated the same way as for scRNA-seq and then resuspended in 49uL of FACS buffer and 1uL of TruStain FcX (BioLegend cat no 422302) and blocked for 10 minutes at room temperature. Following incubation, lyophilized oligo-conjugated antibodies (**Table S2**) were reconstituted in 50uL of FACS buffer and were added to cell suspension and incubated for 30 minutes at 4°C. Then samples were washed 3 times in FACS buffer with centrifugation at 400g for 5 min at 4°C. Single cell suspensions were stained with Calcein AM (Thermo Fisher Scientific C34852) and Propidium Iodide (BioLegend 421301), filtered through a 40-μM cell strainer and immediately brought to the flow cytometry core for sorting. Viable cells were sorted on a BD FACSAria II and selected as being positive for Calcein AM (488 nm excitation and 530/30 bandpass filter) and negative for Propidium Iodide (561 nm excitation and a 610/20 bandpass filter). Doublets were excluded using forward light scatter (FSC) signal area and height as customary. The FACSAria was set up using the 100 um nozzle at 20 PSI pressure, and cells were sorted using a one-drop purity (4-way Purity) mode. Cells were collected in 1.5mL Eppendorf tubes containing DMEM/f12 + 10% FBS before pelleting and counting for 10x Genomics Chromium input.

### CITE-seq library preparation

CITE-seq libraries were prepared as previously described^7, 10^ following 10x Genomics 3’ v3 protocol according to manufacturer’s instructions for cDNA amplification using 0.2 μM of ADT additive primer (5’CCTTGGCACCCGAGAATTCC). The supernatant from the 0.6x SPRI cleanup was saved and purified with two rounds of 2x SPRI, and final product was used as a template to produce ADT libraries. Antibody tag libraries were generated by PCR using Kapa Hifi Master Mix (Kapa Biosciences KK2601), 10 mM 10x Genomics SI-PCR primer (5’AATGATACGGCGACCACCGAGATCTACACTCTTTCCCTACACGACGCTC), and Small RNA RPIx primer (5’CAAGCAGAAGACGGCATACGAGATxxxxxxGTGACTGGAGTTCCTTGGCACCCGAGAATTCCA with xxxxxx denoting one of the four following sequences: CGTGAT, ACATCG, GCCTAA, TGGTCA). Following amplification, Antibody tag libraries were cleaned up with 1.6x SPRI. Subsequently, ADT quality was verified using a DNA high sensitivity assay on an Agilent 2100 bioanalyzer.

### scRNA-seq and CITE-seq data pre-processing

scRNA-seq and CITE-seq FASTQ files were pre-processed by 10x Genomics Cell Ranger 5.0.1 to generate count matrices with quality controls (QCs). Reference version 2020-A with GENCODE v32 annotation was used. For CITE-seq libraries, an additional feature barcode reference was included that contained IDs and sequences of 283 TotalSeq-A antibody-derived tags. Suffix was added to gene symbols when multiple genes with different gene IDs shared the same gene symbol.

### Quality control (QC), normalization, and highly variable gene (HVG) selection

Using the Scanpy package in Python^11^, we first removed any cells expressing less than 200 genes, and then any genes expressed in less than 3 of the remaining cells with functions “pp.filter_cells“ and “pp.filter_genes”. We also annotated the group of mitochondrial genes and computed metrics with function “pp.calculate_qc_metrics”. Cells with total UMI counts over 40,000, or total mitochondrial genes proportion over 30%, or total unique genes over 6,000 were filtered out. Cells that pass QC filtering have gene expression normalized using a two-step procedure: (1) Cell level normalization, in which the UMI count for a given gene in each cell is divided by the total UMI count across all genes in the cell, multiplied by the median total UMI count across all cells, and transformed to a natural log scale. (2) Gene level normalization, in which the cell level normalized values for each gene are standardized by subtracting the mean and dividing by the standard deviation across all cells within the same batch for the given gene. HVGs were selected based on the log-normalized UMI counts using the approach introduced by Stuart and Butler^12^ and implemented in the “pp.highly_variable_genes” function with “batch_key” parameter in the Scanpy package. The selected HVGs were used for downstream clustering analysis.

### Clustering and differential gene and protein expression analysis

CarDEC^13^, a published deep learning tool for scRNA-seq data batch effect correction, gene expression denoising, and clustering of cells, was applied on 15 scRNA-seq samples to get denoised gene expression on the z-score scale. CarDEC_CITE, a modified version of CarDEC for CITE-seq data which preprocesses the gene and protein expression data separately, was utilized on 6 CITE-seq samples, generating denoised feature expression on the z-score scale and identifying 25 clusters.

Marker genes and proteins were detected using “tl.rank_genes_groups” in the Scanpy package for each cluster identified, and the cell types were assigned based on the top features. Major cell lineages were annotated using knowledge of known protein and gene markers for each cell type from previously published flow cytometry and scRNA-seq studies^14, 15^.

### Integrative analysis of scRNA-seq and CITE-seq data

sciPENN is our published deep learning tool for integrative analysis of scRNA-seq and CITE-seq. Leveraging information learned from CITE-seq data, sciPENN can predict protein expression in scRNA-seq and transfer clustering labels from CITE-seq into scRNA-seq data^16^. Using sciPENN, we integrated scRNA-seq and CITE-seq data into a shared latent low-dimensional space, which was further dimension reduced and visualized using functions ”sc.tl.pca”, “sc.pp.neighbors” and “sc.tl.umap” within Scanpy package.

### Sub-clustering of macrophages, SMCs, and Fibroblasts

Among the 25 clusters obtained from the CITE-seq data, cells labeled as Myeloid (clusters 0, 3, 9, 11, 13, 19, 22), SMCs and Fibroblasts (clusters 2, 5, 12, 17, 18) were extracted, respectively. CarDEC was then performed to further cluster and identify the subpopulations. “tl.rank_genes_groups” was used to get marker genes and cell types were assigned based on them.

### Differential gene expression and proportional analysis between symptomatic and asymptomatic plaques

Among 21 samples, 13 are from symptomatic patients, and 8 are from asymptomatic patients. For each cluster, pseudo-bulk RNA-seq analysis on the single-cell data was performed among the symptomatic and asymptomatic groups using DESeq2^17^ package in R. Raw counts were summed per gene per sample for cells in each cluster, pre-filtering was performed to keep genes with at least 10 reads total, standard differential expression analysis was performed using function “DESeq” and results were generated with function “results”. For each cluster, we also calculated the total cell counts proportion per sample, and unpaired two sample t test was performed using “ttest_ind” within scipy.stats package in Python to compare symptomatic and asymptomatic groups.

### Pathway analysis

Cell-type specific differentially expressed (DE) genes, with a Benjamini-Hochberg adjusted p-value of less than 0.01 and log_2_ fold change greater than 1, were uploaded into Ingenuity Pathway Analysis (IPA) software (QIAGEN). IPA analysis reported the −log_10_ p values and Z-scores of canonical pathways and upstream regulators. The highest predicted upregulated canonical pathways and upstream regulators with a −log_10_ p value > 1.3 for each cell population were selected for the respective analysis and visualized with either a heat map or radar plot. Full list of pathways can be found in **Table S4**.

### Gene scoring analysis

To better characterize genes and pathways that are activated or repressed in cell populations, we performed gene scoring analysis in human carotid plaque scRNA-seq data. We first compiled lists of genes involved in multiple cellular functions and pathways, including apoptosis, DNA damage, macrophage efferocytosis, senescence, UPR response, and inflammasome. Cell cycle genes, including 43 S phase genes and 54 G2/M phase genes from Tirosh et al.^18^, were extracted from Seurat package^10^, where gene symbols were updated. UMI counts were normalized to a target total count of 10,000 per cell and then log-transformed. Gene scoring was performed using “scanpy.tl.score_genes” function from Scanpy 1.9.1^11^ with parameters n_bins = 24 and ctrl_size = 100. Cell cycle scoring was performed using “scanpy.tl.score_genes_cell_cycle” function supplied with S phase genes and G2/M phase genes above.

Gene scores were compared between cells from asymptomatic subjects and symptomatic subjects in each of the SMC, modulated SMC, fibroblast, macrophages, and endothelial cell populations. Wilcoxon rank-sum test was performed on each score between asymptomatic and symptomatic subjects in each cluster. P-values were adjusted by Benjamini-Hochberg method. Scores with adjusted P-value <0.05 were considered significantly different between asymptomatic and symptomatic subjects.

To compare gene score between multiple populations, we performed Kruskal-Wallis test across macrophage populations, SMCs and fibroblasts subpopulations. P-values were adjusted by Benjamini-Hochberg method in each set of analyses. For gene scores with adjusted P-value <0.05, Dunn’s test was performed to identify pairs of populations that had significant differences in gene score distribution. P-values from Dunn’s test were also adjusted by the Benjamini-Hochberg method.

### M1 and M2 macrophage signature analysis

To characterize M1- and M2-macrophage functions in macrophage subpopulations, we utilized lists of M1 and M2 signature genes from Martinez et al.^19^. Genes from membrane receptors, cytokines and chemokines, apoptosis-related genes, solute carriers, enzymes, extracellular mediators, and DNA-binding factors were ordered by M1:M2 expression ratio. 54 genes with higher M1-macrophage expression and 43 genes with higher M2-macrophage expression were used to score macrophage subpopulations. Gene score comparison between populations was performed as described above.

### Flow cytometry

Flow cytometry was performed on a frozen cell suspension from a plaque that was previously submitted for scRNA-seq. Following thawing, cells were washed with FACS buffer and centrifuged at 400g for 5 mins. The plaque cells were then blocked with TruStain FcX (BioLegend cat no 101319) for 10 mins at room temperature. Subsequently, BD Brilliant Stain Buffer (BD Biosciences 563794) and the following fluorescently conjugated antibodies were added to the cells for 20 minutes at 4°C: CD45-APC/Cy7 (BioLegend 368515), CD142-APC (BioLegend 365205), CD31-BV421 (BioLegend 303123), CD90-BUV496 (BD Biosciences 741160), and CD29-PerCP-eFluor 710 (Thermo Fisher Scientific 46-0299-41). Cells were then washed 3 times with FACS buffer and viability dyes Calcein AM and Propidium Iodide were added. The fluorescently labeled plaque cells were measured on a Novocyte Penteon and analyzed with FlowJo software.

### Immunohistochemistry of human coronary arteries

The left anterior descending coronary artery was excised from the myocardium and immediately placed into ice-cold RPMI, washed with RPMI to remove any blood, and cut into roughly 5mm sections and embedded in OCT. Blocks were immediately snap-frozen and stored at −80°C. For sectioning, tissue blocks were allowed to equilibrate at −20°C, sectioned into 8um thick sections, and mounted onto Superfrost Plus Microscope Slides.

For immunofluorescence, the sections were fixed with 4% paraformaldehyde for 20 minutes at room temperature. After fixing, the sections were blocked in PBS containing 1% BSA and 10% goat serum for 1 hour. The sections were then subsequently incubated with the following primary antibodies, CD90 (1:200, Abcam ab181469) and Smoothelin (1:1000, Abcam ab219652), overnight at 4°C in a humidified chamber. The next day, the sections were washed with PBS and incubated with the appropriate secondary antibodies and LipidTOX Red (1:200, Thermo Scientific H34476) for 1 hour at room temperature. Finally, the sections were counterstained with DAPI and mounted with Invitrogen Prolong Glass Antifade Mountant (Invitrogen P36982). Images were acquired with a Nikon A1 confocal microscope.

## Results

### High-dimensional multi-omic profiling identifies 25 distinct cell populations in human atherosclerotic plaques

To examine the single-cell transcriptome and cellular immunophenotypes of human atherosclerotic plaques, we performed multi-omic profiling on carotid endarterectomy tissue from 21 patients. Tissue samples were digested enzymatically, and cells from six of these patients were labeled with a panel of 274 CITE-seq antibodies before sequencing. Data were integrated from 88,093 cells across all patients (CITE-seq and scRNA-seq) using our deep learning model sciPENN, which also predicted protein expression in the query scRNA-seq dataset from the CITE-seq reference dataset^16^ (**Fig 1A**). All cells were visualized by uniform manifold approximation and projection (UMAP), revealing 25 distinct cell populations (**Fig 1B**). Utilizing well-established canonical proteins, we identified 2 EC populations and 18 leukocyte populations (**Fig 1C** and **Suppl Fig 1**), including all major immune cell types such as macrophages, T cells, NK cells, B cells, plasma cells, mast cells, dendritic cells, and neutrophils. Within macrophages, we observed 5 subtypes (Macrophage 1-5) that had high, near-ubiquitous expression of major lineage markers such as CD64, CD11c, CD14, and MHCII. Three populations of CD4^+^ T cells (CD4^+^ T Cell 1-3) and 2 populations of CD8^+^ T cells (CD8^+^ T Cell 1 and 2), determined by CD3, CD7, CD2, and CD194 expression, were identified. Cluster 15, known to be phenotypically like T cells, was distinguished by the specific expression of the NK cell markers CD56 and CD16. B Cell 1, B Cell 2, and plasma cells were identified by the expression of CD19 and CD20, and KIT^+^ mast cells were identified by expression of CD117. Two small dendritic cell populations (clusters 19 and 22) that comprised plasmacytoid (pDC) and conventional DCs (cDC) characterized by the expression of CD303 and CD141, respectively, were also identified. We classified cluster 24 as neutrophils, which can be difficult to observe in scRNA-seq data, by the expression of CD15. Similarly, we identified two EC clusters as having specific expression of the proteins CD31, CD34, CD144, and CD49b.

**Figure 1.**
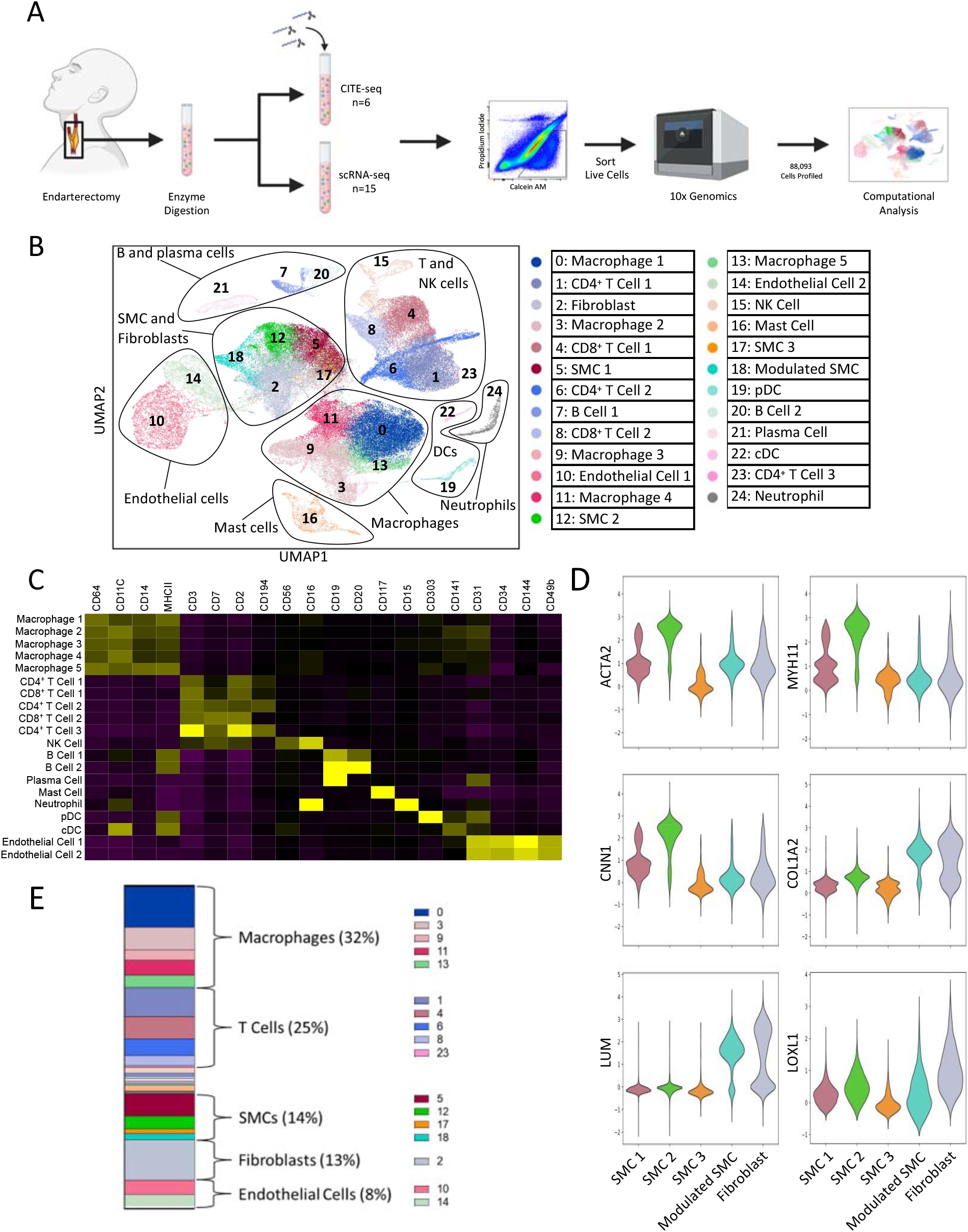
Multiomic analysis of human carotid atherosclerosis identifies 25 distinct cell populations. (A) Experimental design. (B) UMAP visualization of clustering revealed 25 cell populations. (C) Canonical Proteins to identify macrophages, T cells, NK cells, B Cells, Plasma Cells, Mast Cells, Neutrophils, dendritic cells, and endothelial cells. (D) Canonical Genes to identify SMCs and Fibroblasts. (E) Distribution of each major subclass across all samples.

Because SMCs and fibroblasts have not been immunophenotyped extensively, we used canonical gene markers to identify these cell types (**Fig 1D** and **Suppl Fig 2**). To differentiate SMCs and fibroblasts that clustered together by phenotypic similarities, we examined the expression of known SMC genes ACTA2, MYH11, and CNN1, as well as fibroblast-specific genes COL1A2, LUM, and LOXL1. This process revealed that clusters 5 and 12 (SMC 1 and 2) were contractile SMCs with the highest expression of all SMC-related genes, whereas clusters 17 (SMC 3) and 18 (Modulated SMC) had lower expression of SMC genes. Modulated SMC had higher expression of fibroblast genes relative to SMC 1 and 2, indicating that this SMC-derived population has undergone phenotypic modulation. Cluster 2, with high expression of COL1A2, LUM, and LOXL1, encompassed all fibroblasts in our dataset.

Next, we calculated the proportion of each cluster for each of the 21 individual subjects (**Suppl Figure 3A and B**). On aggregate, macrophages comprised 32% of the cells in our dataset, T cells 25%, SMCs 14%, fibroblasts 13%, and ECs 8% (**Fig 1E**). In contrast, other scRNA-seq studies have identified T cells as the dominant cell type in human carotid plaques^8, 9^, perhaps due to differences in digestion and isolation protocols or patient heterogeneity. Leukocytes comprised the largest proportions of cells within our carotid atherosclerosis dataset, consisting mainly of macrophages and T cells.

### Multimodal analysis provides deep understanding of plaque cellular phenotypes

To extend our findings beyond known cell surface protein markers, we used our CITE-seq data to identify the distinct gene and protein signatures for all 25 populations depicted in **Fig. 1B**. To assess the added value of incorporating both RNA and protein information into our analysis, we calculated the gene and protein correlation for every gene-protein pair in our dataset for each individual cluster. As shown in **Fig 2A**, overall, there was a high degree of correspondence between protein and gene expression (**Table S3**), although it is well-established that there can be variability between protein levels and their coding transcripts^20^. In several circumstances, the protein expression contributed critically to the identification of cell types when the RNA was not highly expressed (**Suppl Fig 4**). For example, established neutrophil protein markers CD15 and CD16 were highly expressed and specific although their corresponding genes, *FUT4* and *FCGR3A*, were not detectable.

**Figure 2.**
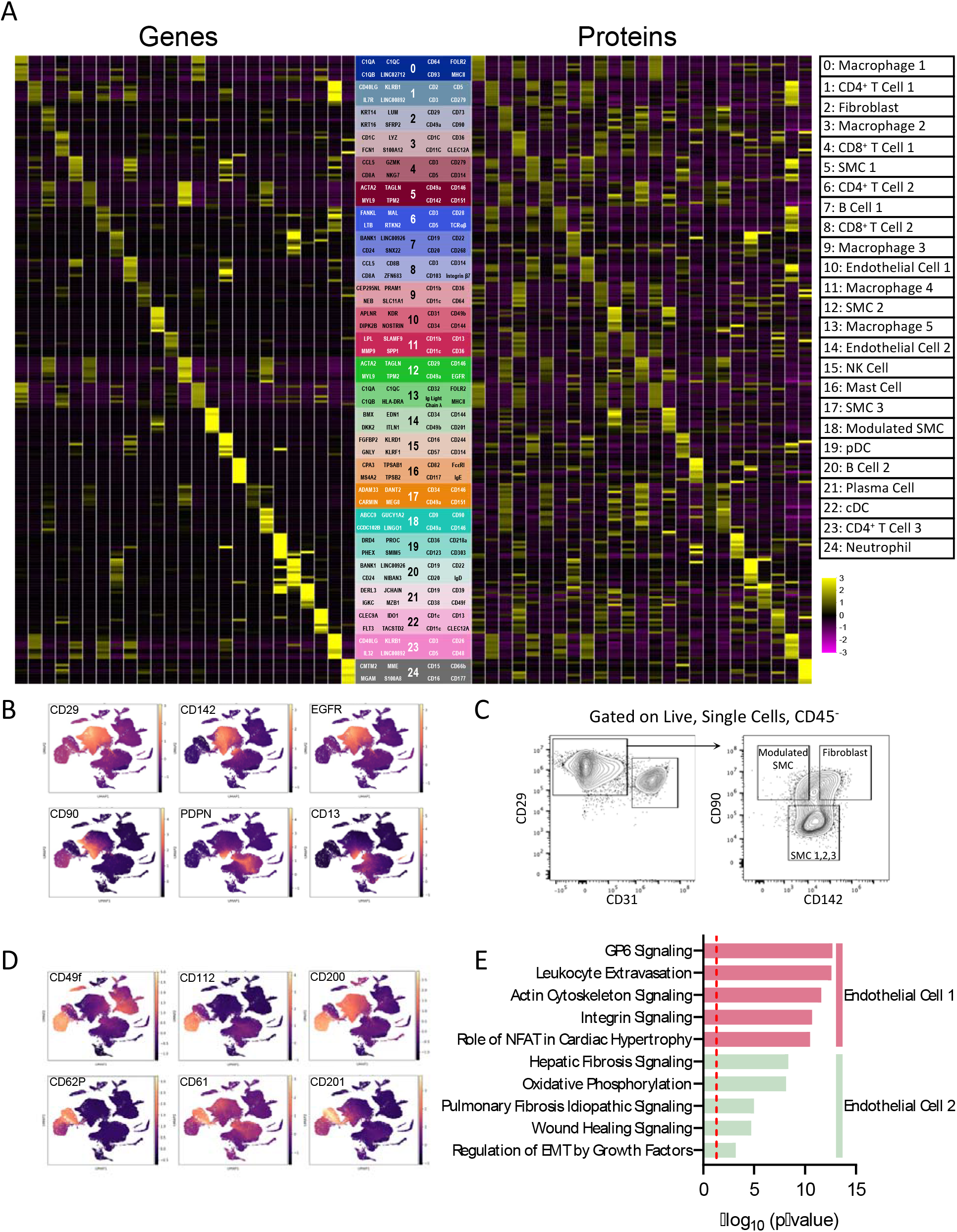
Multimodal biomarkers of cells within carotid atherosclerotic plaques. (A) Heatmap of all cell types identified in carotid atherosclerosis. Markers include top 10 mRNA (Left) and top 10 protein (Right) features identified by differential expression. (B) Feature plots displaying unique protein markers for SMCs (Top) and fibroblasts (bottom). (C) Flow cytometry validation of predicted protein markers for SMC 1-3, modulated SMC, and fibroblasts. (D) Feature plots displaying proteins to differentiate endothelial cell 1 and endothelial cell 2. (E) Pathway analysis comparing endothelial cell 1 and endothelial 2 using genes from differential expression analysis.

Our five macrophage clusters segregated into three major phenotypic classes. Macrophages 1 and 5 had high expression of FOLR2, cadherin 11 and CD93, suggesting roles in inflammation, phagocytosis, and cell adhesion. In contrast, Macrophages 3 and 4 had high expression of CD36 and TREM1, suggesting a role in lipid uptake and metabolism. However, Macrophage 3 uniquely expressed CD35, a receptor for complement activation. Macrophages 2 and 3 shared expression of CLEC12A and CD192 (CCR2), indicating that they are more inflammatory and likely recently recruited from circulation (**Suppl Fig 5A**). Macrophage 4 had the highest expression of genes involved in lipid metabolism such as *ABCA1, LPL, FABP5,* and *APOE*, whereas macrophages 2 and 3 had the highest expression of inflammatory genes such as *IL1B, NLRP3,* and *CCR2* (**Suppl Fig 5B**). Macrophages 1 and 5, notably, had intermediate expression of a substantial proportion of genes involved in both functions. Because there was considerable overlap in the gene expression profile of all macrophage subtypes, we identified which upregulated and downregulated genes were unique to each population (**Suppl Fig 5C**). Using this gene set, we performed pathway analysis to identify in more detail the predicted functional differences between these populations (**Suppl Fig 5D**). This suggested both substantial functional heterogeneity as well as degrees of overlap of plaque macrophages.

CITE-seq profiling was particularly helpful for characterizing SMC, fibroblasts, and ECs using novel protein phenotypic markers. The SMC populations (SMC 1-3) had relatively uniform expression of the proteins CD29, CD142, and EGFR. The modulated SMC population also had relatively uniform expression of CD29 and EGFR but lacked expression of CD142. In contrast, fibroblasts expressed CD90 to a similar degree as the modulated SMC populations, but specifically expressed PDPN and CD13 (**Fig 2B**). Using the combination of these markers, we developed a novel flow cytometry gating strategy to discriminate between SMC 1-3, modulated SMCs, and fibroblasts (**Fig 2C**).

Our clustering also revealed two distinct EC populations (**Fig 1B and C**). To differentiate these, we identified proteins that were expressed uniquely on one but not the other, e.g., CD49f, CD112, and CD200 were expressed on endothelial cell 1 while CD62P, CD61, and CD201 were highly expressed on endothelial cell 2 (**Fig 2D**). To delineate these cell types further, we performed a differential expression analysis followed by pathway analysis using inferred functional differences (**Fig 2E**). This analysis revealed that the endothelial cell 1 population represented activated and inflamed endothelium, suggesting that this cluster promotes leukocyte extravasation. In contrast, the endothelial cell 2 population had upregulation of fibrotic pathways, suggesting that this cluster may be predisposed to undergo endothelial to mesenchymal transition.

### Cerebrovascular events are associated with dysregulation of macrophages as well as specific smooth muscle cell, and endothelial cell subpopulations

UMAP visualization of samples revealed that the majority of clusters overlapped between asymptomatic and symptomatic patient groups (**Fig 3A, 3B**). However, some striking differences were identified, including reductions in the proportions of Macrophage 1, Endothelial Cell 1, SMC 2, and modulated SMC populations, and a modest increase in SMC 3 population in symptomatic patients (**Fig 3C**). The reduction in Macrophage 1, a macrophage population with efferocytotic functions (**Suppl Fig 6A**), suggests that the capacity to clear dead and dying cells is compromised. The decrease in the activated Endothelial Cell 1 population may suggest increased plaque erosion. The decrease in modulated SMCs and SMC 3 subpopulation increase suggests a shift to pro-inflammatory SMCs (**Suppl Fig 6B**). Unlike other studies^8^, we did not see alterations in the proportion of T cell subpopulations (**Suppl Fig 6C**). Contrary to findings in previous publications^8^, differential expression analysis revealed only three significant DE genes associated with clinical status: *S100A8, DDT,* and *PTGS1* (**Suppl Fig 6D**).

**Figure 3.**
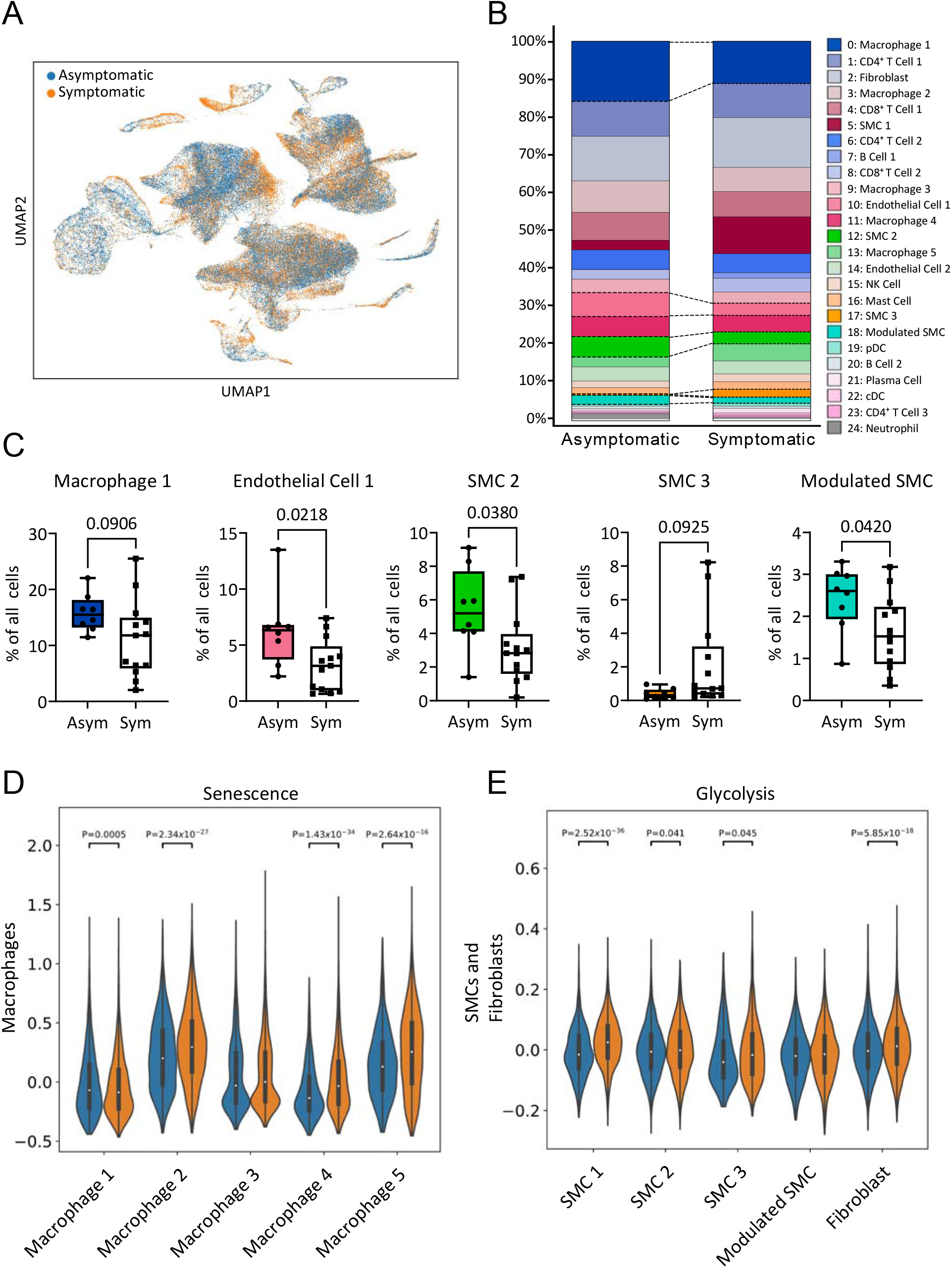
Cerebrovascular events are associated with alterations in the distribution of certain cell populations. (A) UMAP colored by clinical status. (B) Vertical bar graph showing proportion of all clusters by clinical status. (C) Box Plots showing distribution of select cell populations including macrophage 1, endothelial cell 1, SMC 2, SMC 3, and modulated SMC. (D) Gene scoring analysis comparing senescence in macrophage clusters in asymptomatic and symptomatic plaques. (E) Gene scoring analysis comparing glycolysis in SMC and fibroblast clusters in asymptomatic and symptomatic plaques.

To assess biological processes that are associated with clinical events in more depth, we performed a gene scoring analysis for macrophages, SMC, and fibroblasts. Generally, macrophages in symptomatic plaques were more senescent (**Fig 3D**), exhibited a higher level of ER stress through PERK activation, had a higher degree of inflammasome activation, but also were pro-resolving (**Suppl Fig 6E**). Like macrophages, SMCs and fibroblasts from symptomatic plaques were more senescent and PERK was more activated than were those from asymptomatic plaques (**Suppl Fig 6F**). Also, SMCs and fibroblasts were more glycolytic suggesting a metabolic shift in plaques associated with clinical events, as has been noted in plaques with features of plaque instability in mice^21^ (**Fig 3E**).

### Deep sub-clustering reveals diverse macrophages phenotypes in carotid plaques

Initial clustering identified 5 plaque macrophage clusters (Macrophage 1-5) (**Figs. 1 and 2**). To explore cellular diversity more deeply, we performed a sub-clustering analysis on macrophage and DC populations. This revealed 10 cell clusters, 7 macrophage subpopulations, 2 DC clusters, and residual T cells expressing CD3 (**Fig 4A and Suppl Fig 7A and B**). From the initial clustering (**Suppl Fig 7C**), Macrophage 1 mapped to clusters 0 and 4, Macrophage 2 mapped to cluster 1, Macrophage 3 mapped to cluster 3, and Macrophage 4 mapped to clusters 2 and 5. In contrast, Macrophage 5 was distributed throughout the subclusters. Cluster 7 was comprised of cells for every macrophage population in the initial analysis. Clusters 0 and 1 were highly inflammatory, expressing genes *C1Q* and *IL1B*, respectively, and enriched in inflammatory pathways (**Fig 4B**). Clusters 2 and 4 expressed foam cell marker genes, in which cluster 2 expressed *ABCA1, LPL, CD36* and *TREM2* most prominently, whereas cluster 4 expressed *APOE* in addition to inflammatory genes like *CCL18* and *C1Q* genes. Additionally, apoptotic (cluster 3), proliferative (cluster 5), and *ACTA2*^+^ (cluster 7) subpopulations were identified. Cluster 3 showed upregulation of the granzyme A apoptotic pathway (**Fig 4B**), along with high expression of mitochondrial genes (**Suppl Fig 7A and B**). The proliferative subpopulation had specific expression of many proliferation markers including *MKI67, TUBB,* and *STMN1* (**Suppl Fig 7B**). Finally, the *ACTA2*^+^ subpopulation (∼6% of all macrophages) was, notably, characterized by high expression of several SMC-related genes such as *MYOCD, ACTA2, MYH11,* and *CNN1* (**Suppl Fig 7B**), along with protein expression of CD64, CD11c, and MHCII proteins (**Suppl Fig 7D**), suggesting that it could represent phenotypic switching of SMCs to macrophage-like cells. This subpopulation also exhibited upregulation of fibrotic signaling and epithelial-mesenchymal transition pathways (**Fig 4B**).

**Figure 4.**
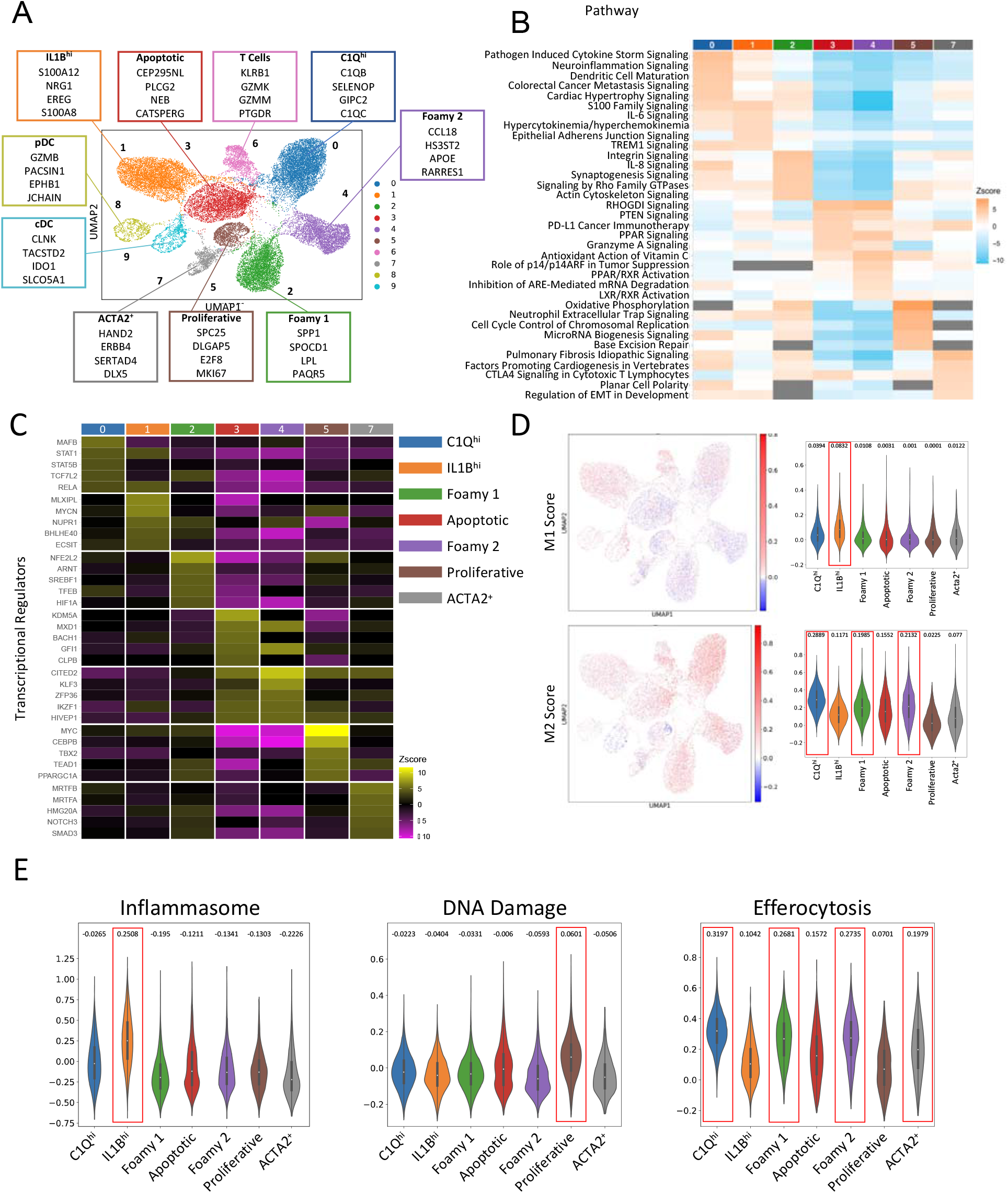
Sub-clustering analysis of myeloid cells reveals further macrophage phenotypic and functional heterogeneity. (A) Sub-clustering analysis of myeloid cells. Macrophage 1-5, pDC, and cDC clusters were selected from the initial clustering analysis, revealing 10 clusters. (B) Heatmap showing top upregulated pathways for each macrophage subpopulation based on top DE genes. (C) Heatmap showing top predicted transcriptional regulators for each macrophage subpopulation. (D) UMAP overlaying expression of common M1 and M2 signatures (left), and violin plots quantifying median expression for each macrophage subpopulation. (E) Gene scoring analysis for inflammasome activation, DNA damage, and efferocytosis across macrophage subpopulations.

To establish regulatory features of each macrophage subpopulation, we conducted an upstream analysis using top DE genes and identified potential transcriptional regulators (**Fig 4C**). The *C1Q*^hi^ (cluster 0) and *IL1B*^hi^ (cluster 1) subpopulations shared regulatory features, like *STAT1* and *RELA*, involved in immune modulation^22, 23^. The proliferative subpopulation (cluster 5) exhibited the most specific regulatory features with high inferred regulation by the oncogene *MYC*. The *ACTA2*^+^ subpopulation showed inferred transcriptional regulation by *MRTFB, MRTFA, NOTCH3*, and *SMAD3*, known to control SMC differentiation^24–27^. To infer M1/M2 canonical functional properties, we tested enrichment of M1- and M2-associated genes^19^ in our dataset (**Fig 4D**). Only *IL1B*^hi^ macrophages showed enrichment in M1 genes. Conversely, the *C1Q*^hi^, foamy 1, foamy 2, and apoptotic subpopulations exhibited strong expression of M2 genes. The proliferative and *ACTA2*^+^ subpopulations did not show a strong M1 or M2 signature. These data align with literature suggesting that the M1/M2 classification inadequately captures the *in vivo* phenotypic diversity of macrophages in atherosclerotic lesions^28^.

Although we did not observe any differences in macrophage subpopulations between asymptomatic and symptomatic plaques (**Suppl Fig 7E**), we evaluated which cell populations had upregulated gene expression for processes known to impact atherosclerosis. To do so, we utilized a list of genes involved in inflammasome activation, DNA damage and efferocytosis to perform gene score analyses for these processes (**Fig 4E**). As expected, the *IL1B*^hi^ subpopulation was the only one with marked inflammasome activation. The proliferative subpopulation had highest expression of genes involved in DNA damage, while each of the *C1Q*^hi^, foamy 1, and foamy 2 subpopulations had evidence for enhanced efferocytotic function.

### Deep sub-clustering reveals multiple SMC and fibroblast subclasses in plaques

In initial clustering we identified 3 SMCs, 1 modulated SMC, and 1 fibroblast population (**Figs. 1 and 2**). This high-level clustering may not reveal the true heterogeneity within these cell types. A recent study that performed snRNA-seq of human aortic tissue identified 7 SMC and 4 fibroblast populations^29^. Therefore, we performed a deep sub-clustering analysis on SMCs and fibroblasts that revealed 10 unique clusters with specific gene expression profiles (**Fig 5A**). Using classical SMC and fibroblast phenotypic markers (**Fig 5B**), we found that clusters 0 and 2 (major SMC 1 and 2) represented the two major classes of contractile SMCs with the highest expression of MYH11 and ACTA2. Two smaller contractile SMC populations, clusters 4 and 6 (minor SMC 1 and 2), were also identified. Minor SMC 1 had moderate expression of MYH11 and TAGLN, but very low expression of other contractile SMC genes. Minor SMC 2 had moderate expression of *MYOCD, MYH11,* and *SMTN*, but low levels of other SMC genes. The most striking population was cluster 5, which had low expression of SMC genes and the specific expression of many genes involved in lipid metabolism, suggesting that this large population may be SMC-derived foam cells (**Suppl Fig 9A)**. This foamy SMC cluster mapped almost exclusively back to the modulated SMC population in the original analysis (**Suppl Fig 9B**). We identified 3 fibroblast populations, clusters 1, 3, and 8, determined by the expression of *BGN, DCN,* and *COL1A1*. Cluster 9, the last cluster identified, represented a fibromyocyte, having the highest expression of *VCAM1* and *LY6E* (**Suppl Fig 9C**).

**Figure 5.**
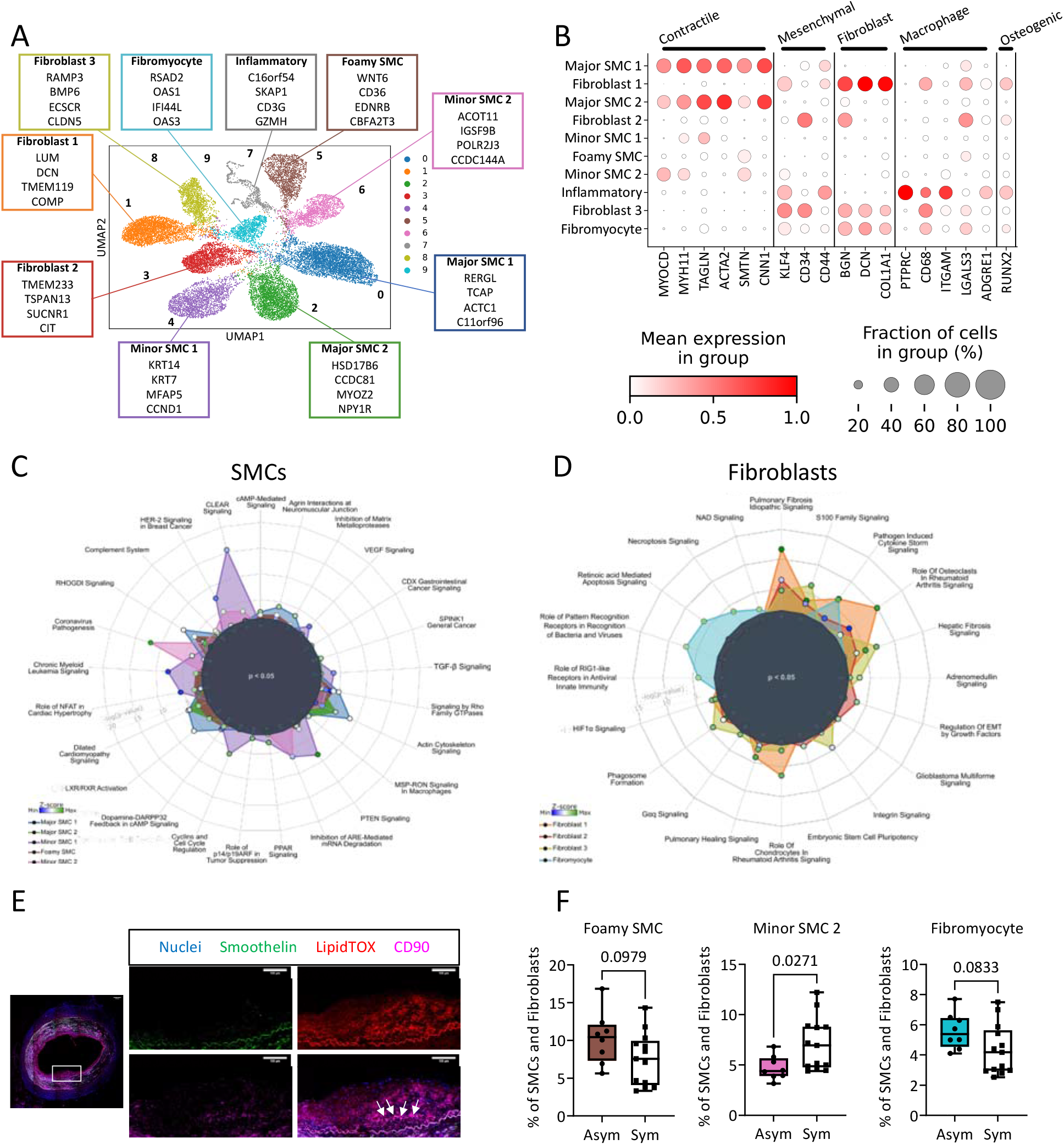
Sub-clustering of SMC and Fibroblasts reveals distinct subpopulations. (A) Sub-clustering analysis of SMC and fibroblasts. SMC 1-3, modulated SMC, and fibroblast clusters were selected from the initial analysis, revealing 10 clusters. (B) Dotplot showing expression of classic SMC genes with corresponding phenotypes. (C) Radar plot showing pathways for SMCs obtained from DE genes. (D) Radar plot showing pathways for fibroblasts obtained from DE genes. (E) Immunohistochemistry staining targeting foamy SMC markers in coronary artery section. DAPI (blue), Smoothelin (green), LipidTOX (red), and CD90 (magenta). White arrows indicate Smoothelin^+^LipidTOX^+^CD90^+^ cells in neointima region of lesion. n=1. (F) Box plots showing distribution of select cell populations including foamy SMC, minor SMC 2, and fibromyocyte comparing asymptomatic and symptomatic plaques.

To identify functional differences in the SMC subpopulations, we performed pathway analyses of SMC and fibroblast sub-clusters. Consistent with annotation as prototypical contractile SMC populations by DE gene analysis, our pathway analyses (**Fig 5C**) revealed that major SMC 1 and 2 were enriched for muscle functions, TGF-β signaling, and actin cytoskeleton signaling. Coincident with downregulation of contractile SMC genes, minor SMC 1 had upregulated pathways involved in mRNA degradation and cell cycle regulation, suggesting that it might be an actively dividing and proliferating subpopulation. Fibroblast clusters had upregulation of several fibrotic pathways, and we found upregulation of necroptotic and apoptotic pathways in the fibromyocytes (**Fig 5D**).

Since our data and those from recent mouse and human studies suggest that many foam cells may be SMC-derived^30, 31^, we used a combination of markers revealed by CITE-seq and functional inference to design an immunostaining protocol to spatially locate foamy SMCs in human coronary artery sections. As noted, the foamy SMC cluster mapped primarily onto the modulated SMC cluster in our initial analysis (**Suppl Fig 9B**), expressed the SMC-specific gene smoothelin (**Fig 5B**), and highly expressed the protein CD90 (**Suppl Fig 9D**). At immunohistochemistry of human coronary artery sections with a combination of smoothelin, CD90, and LipidTOX (to identify foam cells) (**Fig 5E**), we revealed a sub-cluster of neointimal cells that stained for all three markers, validating, and localizing modulated SMC-derived foamy cells in human atherosclerotic lesions.

Given differences in SMC 2, SMC 3, and modulated SMCs by clinical status in our initial clustering (**Fig 3C**), we examined the cell distributions by clinical events in our deep cell type clustering and found subtle decreases in the proportions of foamy SMCs and fibromyocytes, with a significant increase in minor SMC 2 and increased PERK activation in symptomatic plaques (**Fig 5F and Suppl Fig 9E**).

## Discussion

Through the largest and most in-depth scRNA-seq and high-dimensional CITE-seq phenotypic characterization of human carotid atherosclerosis to date, we provide novel insights into the cellular and molecular pathogenesis of atherosclerotic CVD. Our study reveals a comprehensive landscape of 25 cell types, including macrophages and their regulatory features, immunophenotypic and functional characterization of SMCs and fibroblasts, and cell type-specific perturbations associated with cerebrovascular events. We identified 7 macrophage, 3 fibroblast, and 7 SMC subpopulations, including 3 phenotypically modulated or ‘switched’ SMC subpopulations. Each cell population displayed a distinct gene and protein expression signature, reflecting their unique functions. For the first time, we also offer marker panels for immune-isolation and localization of key modulated SMC populations to facilitate further functional and translational studies.

Our work expands on recent advancements in flow cytometry and CyTOF that have improved our understanding of atherosclerosis by revealing the diversity of cell types in this disease^8, 32, 33^ and also contributes beyond those made to date by single-cell genomic technologies, as exemplified in the work by Fernandez et al.^8^ and Depuydt et al.^9^. These studies have enhanced our knowledge of cellular heterogeneity and regulatory networks in atherosclerosis^8, 9, 26, 34–43^. Fernandez et al.^8^ used CyTOF, scRNA-seq, and selective CITE-seq to investigate immune cell diversity in human carotid atherosclerosis, but their study lacked nonimmune cells and only single-cell profiled 6 patients. Depuydt et al.^9^ expanded the scRNA-seq analysis to 18 patients, including immune and nonimmune cells, and performed scATAC-seq to identify transcriptional regulators. However, both studies lacked detailed phenotypic characterization of cell types within lesions, which is critical for understanding their roles in the disease.

Despite extensive evidence in mouse and human atherosclerosis that SMC, SMC-derived cells, and fibroblasts comprise the majority of cells within atherosclerotic lesions^53^, single-cell profiling studies in humans have focused primarily on leukocytes. To address this gap, we performed a comprehensive characterization of SMCs and fibroblasts in human lesions. Our analysis identified unique surface proteins, including CD29 and EGFR, that are expressed universally across these cell types, while CD90 was specific to modulated SMCs and fibroblasts, and CD142 was not expressed on the modulated SMC population. These findings enable isolation of these cell types by flow cytometry and determination of the spatial location of each cell type within atherosclerotic tissue, thereby providing critical insight into their functions and potential cell-cell interactions. Sub-clustering analysis revealed previously unknown heterogeneity, including two major contractile SMC and two modulated SMC subpopulations (foamy and fibromyocyte). Mouse models suggest that fibromyocytes contribute to plaque stabilization^35^, while the role of SMC-derived foam cells remains unclear. Our preliminary evidence suggests that SMC-derived foam cells are localized to the neointima of advanced coronary lesions, not the fibrous cap, shedding light on their possible functions and lesion domain interactions in atherosclerotic lesion subdomains.

Our extensive plaque characterization suggests that macrophages are the predominant leukocyte population, contrary to findings from Fernandez et al. and Depuydt et al., who identified T cells as the most abundant immune cell type in lesions. Previous scRNA-seq studies of mouse and human lesions indicated a higher prevalence of myeloid cells than T cells^35, 38, 39^, highlighting the need for standardization of sample processing, analytical methods, and detailed cross-species comparisons. Using protein and gene expression, we identified distinct phenotypic, functional, and activation states of plaque macrophages. Sub-clustering analysis confirmed two proinflammatory macrophage subsets (*IL1B*^hi^ and *C1Q*^hi^) with enriched inflammatory pathways, consistent with other single-cell studies in different disease contexts^44^. The *IL1B*^hi^ subset exhibited the most distinct inflammatory gene and protein expression profile, including the genes *IL1B, NLRP3, CCR2*, and the protein CD192. This suggests that *IL1B* targeting with canakinumab, which reduced CVD events in the CANTOS trial^45^, may act preferentially on these macrophages. Of further translational relevance, our recent work in mice with collaborators suggests that inhibiting *IL1B* in individuals with clonal hematopoiesis (CH) could effectively reduce CH-driven CVD risk by targeting inflammatory macrophages^46^.

Our macrophage findings support, in part, a recent transformative study in mice that identified nonfoamy macrophages as having a distinct proinflammatory transcriptome and suggested that lipid accumulation in macrophages may not be a driver of plaque inflammation and vulnerability^36^. Yet our data suggest complex functions for macrophage foam cell sub-types. We discovered two distinct foamy macrophage types in lesions. Foamy 1 had a classic foam cell phenotype with enriched lipid metabolism genes (*ABCA1, LPL, FABP5*) and high CD36 protein expression. This population predominantly expressed TREM2, which is proposed to protect against atherosclerosis by facilitating cholesterol metabolism through inducing expression of *APOE, LPL,* and *LIPA*, while simultaneously reducing inflammation^47^. However, in potential conflict with this model, a recent preprint revealed that macrophage-specific deletion of Trem2 in mice reduced atherosclerosis progression and plaque burden^48^ suggesting that TREM2 may be atherogenic at least under certain circumstances. These studies highlight the critical need for additional research in this area e.g., to determine if Trem2 agonism or antagonism and under what circumstances is a viable approach for preventing atherosclerosis. In this context, our Foamy 2 macrophage population expressed TREM2 and certain foam cell genes (*APOE*) as well as *C1Q* genes, indicating an intermediate state between Foamy 1 and inflammatory C1Q^hi^ populations. In fact, we identified more than one cluster in our initial analysis (Macrophage 1 and 5) that had intermediate expression of genes involved in both inflammation and lipid metabolism. Further research is needed to understand the functions of these foamy-inflammatory macrophage types and their impact on disease progression and clinical events.

We found a macrophage subpopulation expressing proliferative genes, like the stem-like macrophages identified in murine atherosclerosis through fate-mapping^49^. Macrophage proliferation is prominent in advanced stage atherosclerosis in mice^50^ and can also occur distinctly during lipid-induced regression and plaque stabilization^51^. Thus, this proliferative population may be responsible for local macrophage proliferation in advanced human atherosclerosis or may act as a transitional cell state giving rise to various phenotypes, including macrophages involved in plaque regression and stabilization. These findings highlight the diversity and complexity of macrophage subtypes within plaques^52^, emphasizing opportunities but significant uncertainties in targeting specific macrophage populations with precision therapeutics.

Our study supports the concept of marked cell type transitions in human atherosclerosis. We identified a small (∼6% of all macrophages) but distinct population of macrophages that expressed SMC-specific genes and displayed a fibrotic pathway phenotype regulated by known SMC transcription factors^26, 54^. Interestingly, these macrophages also expressed classic macrophage proteins, which has been observed previously in advanced human coronary atherosclerosis^30^, suggesting they are of SMC origin. This finding aligns with previous mouse studies showing SMC plasticity during atherosclerosis progression, including SMC-derived macrophages, fibroblasts, osteogenic cells as well as foam cells^31, 34, 35, 39^. Additionally, we identified a substantial population of SMC and likely SMC-derived cells that had a foam cell gene signature, with specific expression of genes involved in lipid metabolism, agreeing with recent studies suggesting that a large portion of foam cells within atherosclerotic plaques may be of SMC origin^30, 31^. We also observed two distinct EC populations, with one showing upregulated fibrotic pathway genes suggestive of endothelial-to-mesenchymal transition. Our findings provide human evidence that supports recent studies in mice, highlighting the heterogeneity and transitions of ECs during atherosclerosis^34^. Importantly, this EC pathophysiological process has been linked to increased disease severity in mouse models of atherosclerosis and in human coronary disease^55, 56^.

We found that specific cell-type alterations may associate with symptomatic clinical events. Our clinical analyses, however, may have limitations due to small sample sizes, patient selection biases, and time delays between cerebrovascular events and surgeries. Despite limitations, we observed decreases in efferocytotic macrophages, activated ECs, contractile SMCs, and foamy modulated SMCs, along with an increase in inflammatory SMCs in symptomatic plaques. These alterations indicate a loss of efferocytotic macrophages, erosion of the EC monolayer, and a shift towards an inflammatory SMC/SMC-derived state, in plaque vulnerability^2^. Additionally, macrophages in symptomatic plaques showed increased senescence, also reported to associate with plaque vulnerability^57, 58^. SMCs in symptomatic plaques exhibited an increased glycolytic gene signature, extending prior findings of altered metabolism in plaques overall of high-risk patients^59^. As a corollary, alterations in bioenergetic mechanisms in mouse models of atherosclerosis have been found to control SMC differentiation and to be associated with features of plaque instability^21, 60^. These clinical associations provide the foundation for pre-clinical and therapeutic studies targeting specific cell types and molecular programs to improve outcomes in atherosclerotic CVD, particularly in high-risk patients unresponsive to lipid-lowering therapy.

Although our dataset provides extensive insights into human carotid atherosclerosis, its sample size is still limited and prone to technical biases and artifacts. Findings need to be further corroborated in human coronary arteries and for cardiac events. To gain more confidence in identifying causal alterations associated with clinical CVD events, it is also crucial to increase the number of patient samples and diversity, and to utilize advanced computational and integrative genomics methods and large-scale human genetics. These integrative studies can help identify specific cell types and molecular programs that play a causal role, enabling mechanism-based translation for clinical and therapeutic applications.

In summary, we present the most advanced in-depth multimodal atlas of advanced human carotid atherosclerosis, reveal a vast cell-type heterogeneity of macrophages, SMCs, and fibroblasts, and identify cell type-specific alterations associated with clinical events. We characterized SMC and fibroblasts in unprecedented depth, revealing 3 populations of phenotypically modulated SMCs, including a foamy population that resides in the deep intima of the developing lesion. Further, we have uncovered complex macrophage subpopulations with distinct gene, protein, and transcriptional regulatory features. These data are foundational for future studies mapping CVD susceptible loci to specific cell types, with the goal of identifying new CVD targets specific for certain cell populations fundamental to the progression and regression of disease. Such integrative approaches provide the analytical and conceptional framework for the discovery of novel therapeutics that target specific cell populations in CVD.

## Data Availability

De-identified single cell RNA sequencing, CITE-seq, clinical, and demographic datasets from the n=22 consented individuals described in this study (distributed among 21 carotid and 1 coronary artery specimens) will be deposited in NCBI GEO for public access at manuscript publication.

## Acknowledgements

The scRNA-seq and CITE-seq (10x Genomics) was performed in the JP Sulzberger Columbia Genome Center, supported in part through the National Institutes of Health/National Cancer Institute Cancer Center Support Grant P30CA013696, and used the Genomics and High Throughput Screening Shared Resource. Flow cytometry experiments described in this article were performed in the Columbia Stem Cell Initiative Flow Cytometry core facility at Columbia University Irving Medical Center. Confocal images were collected in the Confocal and Specialized Microscopy Shared Resource of the Herbert Irving Comprehensive Cancer Center at Columbia University, supported by National Institutes of Health (NIH) grant no. P30 CA013696 (National Cancer Institute).

